# The impact of knowledge, attitudes, and practices on hypertension Control and associated Comorbidities among workers of a beverage company in Bangladesh

**DOI:** 10.1101/2024.11.19.24317597

**Authors:** Abu Sayeed Md. Saleh, Monjur Rahman, S M Saidur Rahman Mashreky

## Abstract

Hypertension is well known globally as a major health issue because of its high magnitude and further complications of cardiovascular and kidney diseases. Bangladesh has witnessed a gradual epidemiological transition where communicable diseases especially the chronic ones are dominating including the rural area where people have least access to the health care services. As is evident from aforementioned complications, hypertension if not well managed, equally has dire consequences hence calls early diagnosis and intervention. Consequently, hypertension prevalence in low-income countries rises due to certain lifestyle aspects; for instance, Afghanistan. In regard to this, Knowledge, Attitude, and Practice (KAP) questionnaires facilitate in assessing gaps in the approach toward managing hypertension. The same issues are seen in Benin: China, Egypt: Ethiopia, Ghana: India: Iran, Lebanon, Malaysia: and Mongolia where the government acknowledges that solutions should be specific to increase awareness, act on lifestyle factors and therapy compliance. Thus, management of hypertension must address general approaches to reducing the burden of hypertension and its effects on the affected individuals and healthcare facilities across the world.

## Introduction

Hypertension remains a leading and complex worldwide health problem that is expressed by high incidence of cardiovascular and renal diseases [1]. In Bangladesh there is an epidemiological transition from acute communicable diseases to chronic non-communicable diseases including hypertension; however, hypertensive individuals are more in rural areas who are poorly served by health facilities [1]. Hypertension if not well managed will result in serious conditions such as heart attack, stroke, and kidney failure hence the need to screen for it early [2]. Again, high blood pressure is not a thing of the past in the low-income countries like Afghanistan because influences like obesity, lack of exercise, and high intake of salty foods persist [2]. KAP surveys gauge knowledge and perception towards health issues with the view of establishing deficits of health systems [2]. In the same manner, hypertension in Benin is quite rampant, and there is a poor level of knowledge in patients and low compliance with differential controlling factors [2]. China now has a growing population of, especially the elder group, suffering from hypertension and therefore, knowledge, awareness, and behavior concerning hypertension play an essential role [4].

A different approach of management of hypertension was adopted in Egypt that incorporates biological, psychological, social and culture aspects [5]. Ethiopia and other low- and middle-income countries have also encountered increasing rates of hypertension, and thus, there is the need to increase the patient’s awareness and compliance with management alterations [6]. Hypertension and diabetes are common conditions in Ghana but these diseases are mostly concealed, therefore formulating knowledge, attitude and practice of the general populace matters. Hypertension is a major cause of morbidity and mortality in the large population of India, and hence, there is a need for programs to change the knowledge and subsequent behavior related to the condition [8-11]. Overall, hypertension prevalence in Iran is high, and studies involving KAP questionnaires indicated some shortcomings in the practices regarding hypertension [27, 29, 30]. In Lebanon, high rates of hypertension have been recorded, however, the respective patients display a poor level of awareness and symptom identification, therefore Strengthening KAP is required [13].

Like any country, the management of hypertension in the Malaysian population has its barriers that need measures to overcome and enhance knowledge and mitigate the effects of improved lifestyles contributing to hypertension [14]. Hence, Mongolia with very high levels of poorly controlled hypertension, aims at increasing hypertension knowledge among health care providers in order to improve management results [15].

## Materials and Methods

### Ethical Considerations

Ethical (IRB) approval was obtained from the ethical committee of North South University (Reference Number: 2024/OR-NSU/IRB/0403 prior to the conducting of this study. Each questionnaire began with an Informed consent form covering the background, objectives and Procedures regarding the study. Assurance was provided to the respondents of the completely voluntary nature of their participation and the freedom to leave the study participation anytime they want. All information related to the respondents was kept confidential and all filled forms were stored securely. This study was strictly conducted based on the principles formulated by the 18th World Medical Assembly declaration of Helsinki (Helsinki, 1964) and all applicable amendments.

### Study Design and Study Population

This descriptive type of cross-sectional study was conducted among employees of a Bangladeshi beverage company in the district of Dhaka to assess the Knowledge, Attitude and Practice (KAP) towards hypertension. Eligible respondents were the management and non management staff of a beverage company, > 18 years old, had willingness to participate in the study and who had given informed consent. Cochrane formula (Z = 1.96 and alpha error = 0.05) and Knowledge level (66.7%) from a previous study was used and the required sample size counted to be 340.

### Tools and Procedures of Data Collection

#### Questionnaire

A face-to-face structured questionnaire investigation was carried out for all participants at the beverage company premises. The questionnaire consisted of 30 questions covering the demographic features (age, gender, marital status, socioeconomic status) and KAP on hypertension. The survey took approximately 15 minutes per participant to complete.

The questionnaires for this study were developed based on literature review and adjusted based on the requirements demanded by the study. For the aspects of clarity and relevance of the instrument, the questionnaire was pre-tested with a sample of 20 employees working in the specific beverage company to check its face validity. Information related to patients’ age, gender, education level, and SES (Socioeconomic Status) were gathered. SES was determined according to the monthly income of the participants: Classified based on price range they start from 10000 BDT (low), 10000-40000 BDT (middle), and > 40000 BDT (high). The study was approved by the North South University’s Institutional Review Board for the conduct of research on human participants (#2024/OR-NSU/IRB/0403) and followed the revised Declaration of Helsinki. The survey details were explained to the respondents, and each of them signed informed consent. Privacy was maintained and no data gathered from the interviews was revealed to any person who had no right to know it.

#### BP measurement

Each participant was told to refrain from cigarette smoking, coffee/tea, and exercise for at least 30 min before coming to the specified study location (Doctor’s chamber) and allowed to rest for at least 15 min before their BP measurements were taken, using a sphygmomanometer (Mercury). Each participant contributed to 3 BP readings at 2 minute intervals in a sitting position. A trained primary health care personnel carried out the BP measurements. The average of the last two of the three readings was used for analysis.

#### BMI measurement

The weight of each participant was measured using a standard weight machine and height was measured using a standard stadiometer. The BMI was measured with the following formula (weight in kg/ height in meter squared)

#### Definition

Hypertension was defined as systolic BP (SBP) ≥ 140 mm Hg and/or diastolic BP (DBP) ≥ 90 mm Hg, and/or self-reported current treatment. Participants with controlled hypertension were classified as subjects with systolic BP (SBP) b140 mm Hg and diastolic BP (DBP) b 90 mm Hg. Current use of antihypertensives and/or lifestyle change was defined as treatment of hypertension.

#### Statistical analysis

All data entry and analysis was done with Statistical Package for the Social Science version 26 (SPSS). Using the requirements of the method, descriptive statistics were offered for all the variables in the model. Sampled participants were 340 of which demographic profiles of the respondents were assessed within the domain of hypertension within the context of knowledge attitude and practice (KAP). The Chi-square (χ2) test was applied to compare correlations between variables Though, a p-value of <0. 05 considered statistically significant.

Lack of awareness about the cases of hypertension was estimated by summing the responses to the questions 1-7 where “Agree” responses were offered as well as the responses to the questions 8 & 9 where “Disagree” responses were offered. The subjects’ incorrect knowledge was established by adding the number of the responses which were marked as “Neutral” or “Disagree” to the questions number 1-7 and the responses marked as “Neutral” or “Agree” to the questions number 8 and 9.

Positive attitudes on hypertension were determined by adding the scores for question 10, that affirms that hypertension causes an increase in blood pressure; 11, that affirms that hypertension is an illness; 13 that affirms self-treatment of hypertension is dangerous; and 14 that affirms regular control of hypertension is important, while excluding the score of question 12, which affirms that hypertension is not a chronic disease. Self-reported negative attitudes were determined by adding the “Neutral” and “Disagree” responses to 10, 11, 13, and 14 and the “Agree” response to question 12.

In the evaluation of the level of hypertension management, the total of the “Agree” options in questions 18, 19 and 20 were considered as good practice scores. Practices that were perceived as poor were obtained by totaling the average score of ‘Agree’ and ‘Neutral’ to the following questions; fifteen, sixteen, seventeen, twenty–one, twenty–two and twenty–three. These assessments gave information to the lifestyle behaviors with hypertension in the studied population.

In case of Knowledge, Attitude and Practice, ‘Don’t Know’ responses have been included into Incorrect knowledge, Negative Attitude and Poor Practice respectively.

## Result

### Demographics and Participant Characteristics

Socio-demographic information for the 340 respondents that returned questionnaires are shown in Table 1. The highest proportion of the participants fell within the 30-39 age bracket, at 36.8%, followed by the 20-29 age bracket, which consisted of 35.6%, while 21.8% were between 40-49 years old. A smaller proportion of 5.9% belonged to the age group of 50-59 years. In the sample, the majority were male, at 97.6% of the total number of participants, whereas female participants accounted for about 2.4% of the number. The educational background revealed that 47.4% had completed their higher secondary education (HSC), followed by 45.3% who were graduate degree holders. Only a few of the participants reported having primary or secondary education: 3.5 and 3.8%, respectively. Most of the participants in this study were married, with 97.6% stating their marital status as married and only 2.4% reported as single. Socio-economic status was reasonably well spread, with 27.9% classified as low SES, 36.5% as middle SES and 35.6 % as high SES.

**Table 1.** Socio-demographic characteristics of the respondents in the survey (N = 340)

| Variables | N (%) |
| --- | --- |
| **Age** |  |
| 20 - 29 years | 121 (35.6) |
| 30 - 39 years | 125 (36.8) |
| 40 - 49 years | 74 (21.8) |
| 50 - 59 years | 20 (5.9) |
| **Gender** |  |
| Male | 332 (97.6) |
| Female | 8 (2.4) |
| **Educational Status** |  |
| *Primary School* | 12 (3.5) |
| *Secondary Education (SSC)* | 13 (3.8) |
| *Higher Secondary Education (HSC)* | 161 (47.4) |
| *Graduate* | 154 (45.3) |
| **Marital Status** |  |
| *Single* | 8 (2.4) |
| *Married* | 332 (97.6) |
| **Socioeconomic Status (SES)** |  |
| *Low* | 95 (27.9) |
| *Middle* | 124 (36.5) |
| *High* | 121 (35.6) |

### Knowledge, Attitudes, and Practices Relating to Hypertension

From Table 2 are the outline details on the respondents’ knowledge, attitudes and practices as they relate to hypertension. Knowledge in respect to hypertension was relatively poor, with just 31.5% of the participants answering questions related to the basics about hypertension correctly, while 68.5% had misinformation. Precisely, 42.9% correctly identified hypertension as high blood pressure that is always elevated, while 48.8% were uncertain. To the statement that normal blood pressure is 120/80 mmHg, 30.9% agreed while 35.9% were not sure, and 33.2% disagreed. Only 29.1% recognized that a healthy weight can lower blood pressure, while 37.9% were uncertain, and 32.9% disagreed. Similarly, only 29.1% agreed that a reduction in the intake of salt helps an individual manage and maintain their blood pressure level, while 37.1% were not sure, and 33.8% disagreed. It is helpful to point out that while regular exercise was found helpful by 38.5%, 40.0% remained unsure and 21.5% disagreed. Hypertension was not caused by stress according to 55.0% of the respondents, and 24.7% were undecided. Hardly anyone was aware of the severe health risks of hypertension since only 24.1% regarded the disease as a serious health hazard; 37.6% were not sure and 38.2% disagreed. Regarding drug adherence, 17.1% agreed that it was not necessary to take medication when one was asymptomatic; 37.6% were undecided, and 45.3% disagreed.

**Table 2.** Knowledge, attitudes and practices regarding hypertension.

| Questions |  | Agree<br>n (%) | Don't<br>know<br>n (%) | Disagree<br>n (%) |
| --- | --- | --- | --- | --- |
| **Knowledge** |  |  |  |  |
| *Q1* | *Hypertension refers to a condition with consistently high blood pressure.* | 146(42.9) | 166, (48.8) | 28 (8.2) |
| *Q2* | *A normal blood pressure measurement is 120/80 mmHg.* | 105(30.9) | 122 (35.9) | 113(33.2) |
| *Q3* | *A healthy weight can help lower blood pressure.* | 99(29.1) | 129 (37.9) | 112(32.9) |
| *Q4* | *Eating a diet low in salt can help control blood pressure.* | 99 (29.1) | 126 (37.1) | 115 (33.8) |
| *Q5* | *Regular physical activity can help lower blood pressure.* | 131 (38.5) | 136 (40.0) | 73 (21.5) |
| *Q6* | *Stress can contribute to high blood pressure.* | 69 (20.3) | 84(24.7) | 187 (55.0) |
| *Q7* | *High blood pressure (hypertension) can cause serious health problems like heart disease, stroke, and kidney failure.* | 82(24.1) | 128(37.6) | 130 (38.2) |
| *Q8* | *If you feel well, you don't need to take medication for high blood pressure.* | 58 (17.1) | 128 (37.6) | 154 (45.3) |
| *Q9* | *There is a cure for high blood pressure.* | 110(32.4) | 127 (37.4) | 103 (30.3) |
| **Attitude** |  |  |  |  |
| *Q10* | *It is important to have my blood pressure checked regularly.* | 75(22.1) | 133 (39.1) | 132(38.8) |
| *Q11* | *Making healthy lifestyle changes is an effective way to manage high blood pressure.* | 102(30.0) | 121 (35.6) | 117 (34.4) |
| *Q12* | *Taking medication for high blood pressure is inconvenient.* | 78(22.9) | 140(41.2) | 122(35.9) |
| *Q13* | *I am confident that I can control my blood pressure.* | 86 (25.83) | 123 (36.2) | 131 (38.5) |
| *Q14* | *It is important to follow a doctor's recommendations for managing high blood pressure.* | 80 (23.5) | 136 (40.0) | 124(36.5) |
| **Practice** |  |  |  |  |
| *Q15* | *I currently smoke tobacco products, such as cigarettes, cigars, or pipes.* | 154 (45.3) | 131 (38.5) | 55 (16.2) |
| *Q16* | *I smoke tobacco products daily.* | 166 (48.8) | 128 (37.6) | 46 (13.5) |
| *Q17* | *During the past 30 days, someone has smoked in my surroundings or among my peers.* | 157 (46.2) | 145(42.6) | 38 (11.2) |
| *Q18* | *I eat fruit at least 3 days a week.* | 104 (30.6) | 120 (35.3) | 116(34.1) |
| *Q19* | *I eat vegetables at least 3 days a week.* | 109 (32.1) | 122 (35.9) | 109 (32.1) |
| *Q20* | *I spend at least 20 minutes walking or bicycling for travel on a typical day.* | 98 (28.8) | 129 (37.9) | 113(33.2) |
| *Q21* | *I usually spend more than 3 hours sitting or reclining on a typical day.* | 114 (33.5) | 120 (35.3) | 106(31.2) |
| *Q22* | *I often add salt or a salty sauce to my food right before or as I am eating it.* | 90(26.5) | 140 (41.2) | 110 (32.4) |
| *Q23* | *Salt, salty seasoning, or a salty sauce is often added in cooking or preparing foods in my household.* | 91 (26.8) | 137 (40.3) | 112(32.9) |

Regarding the management of hypertension, participants’ attitudes were as follows: the necessity of checking their blood pressure regularly was believed by 22.1%, 39.1% were undecided, and 38.8% did not believe in its necessity. Only 30% of them regarded lifestyle modifications as an effective treatment, although 35.6% were undecided and 34.4% did not believe in it. Concerning medication, 22.9% considered it a bother to use, while 41.2% were undecided. As for response efficacy, 25.8% of the participants were certain of their competency in controlling their blood pressure, though 36.2% were undecided and 38.5% were not sure.

The participants’ practices about hypertension and lifestyle factors were highly varied in terms of health behaviors. Almost half, 45.3%, reported smoking, of which 48.8% smoked every day. Consumption of fruits and vegetables at least three days a week was reported by 30.6% and 32.1%, respectively. Only 28.8% performed some form of physical activity such as walking or bicycling for at least 20 minutes per day while 33.2% disagreed with the statement. It was also a common activity to spend more than three hours per day sitting or reclining, with 33.5% responding positively. In addition, frequent addition of salt at a table was done by 26.5%, while 26.8% also reported frequent use of salty seasoning during food preparation.

Knowledge, Attitude, and Practice Scores The results represented in Table 3 give insights into the general knowledge, attitude, and practices that participants have about hypertension. As many as 86.2% of the respondents wrongly perceived hypertension, whereas only 13.8% had the right knowledge. This indicates a huge gap in knowledge, as most of them were unaware of the actual facts related to the nature of hypertension, its causation, and how to manage it effectively.

**Table 3.** Knowledge, attitudes and practice score.

| **Questions** | **N (%)** |
| --- | --- |
| **Knowledge Score** |  |
| *Correct Knowledge* | 47 (13.8) |
| *Incorrect Knowledge* | 293 (86.2) |
| **Attitude Score** |  |
| *Positive Attitude* | 75 (22.1) |
| *Negative Attitude* | 265 (77.9) |
| **Practice Score** |  |
| *Good Practice* | 79 (23.2) |
| *Poor Practice* | 261 (76.8) |

Regarding attitudes, the majority expressed negative attitudes toward the management of hypertension, 77.9%, while only a small minority revealed a positive outlook on the importance of hypertension management, 22.1%. This means that most of the respondents either did not view the issue as important or were unmotivated in their actions, which could have an effect on their intention in terms of preventative or health-enhancing behavior.

In analyzing lifestyle practices, it was noted that 76.8% of the respondents had poor lifestyle practices in managing hypertension, while 23.2% had habits that were beneficial to manage hypertension. A result of this nature indicates that most participants may fail to adhere to recommended lifestyle practices such as healthy eating, regular physical activity, or other health-enhancing behaviors that would aid in the control of hypertension.

### Socio-Demographic Factors Associated with KAP

Chi-square analysis was done in Table 4 to find the association of socio-demographic factors with the levels of knowledge, attitude, and practice regarding hypertension. Most of the socio-demographic variables did not show statistical significance with KAP scores, except a few.

**TABLE 4:**
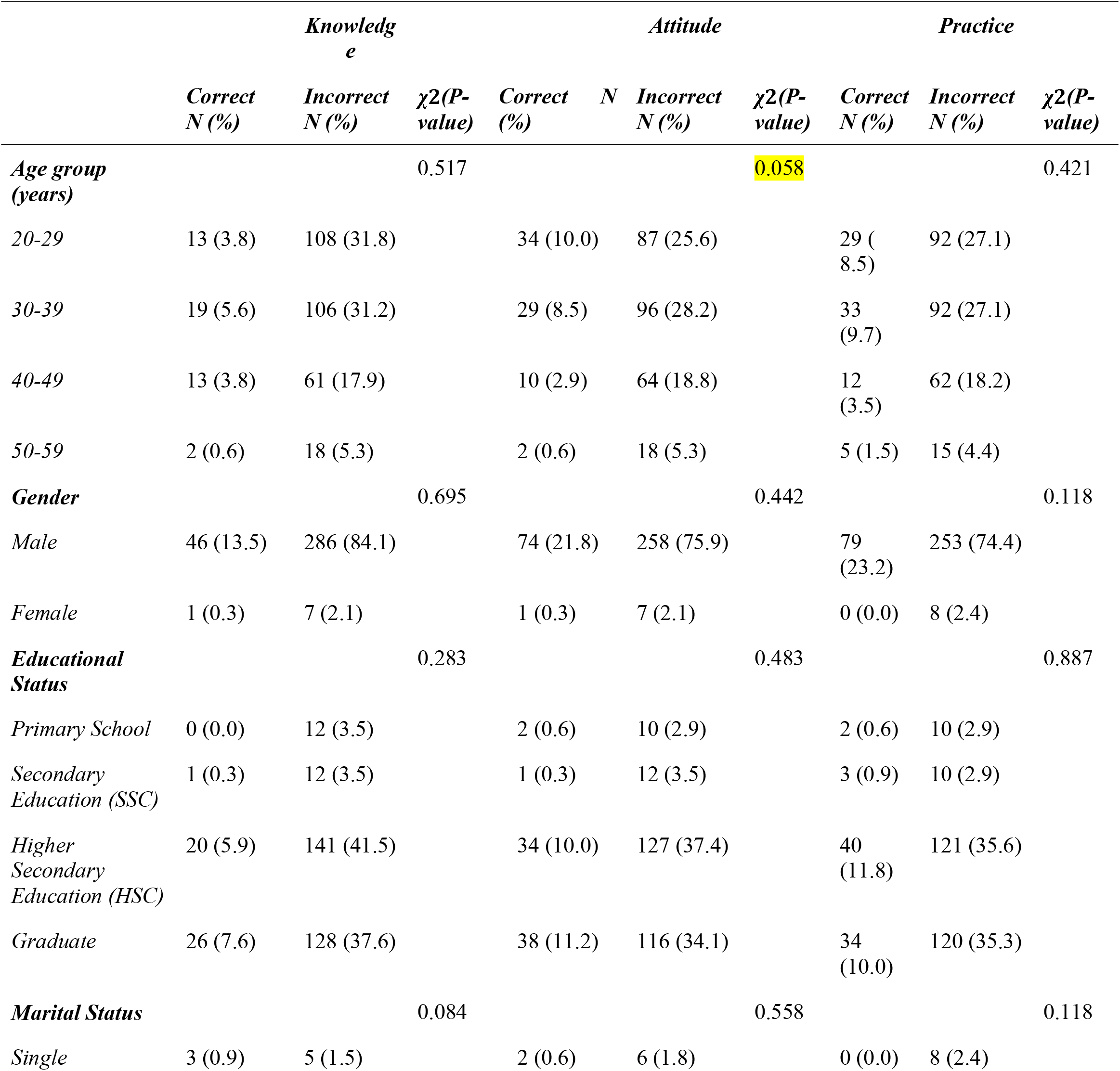
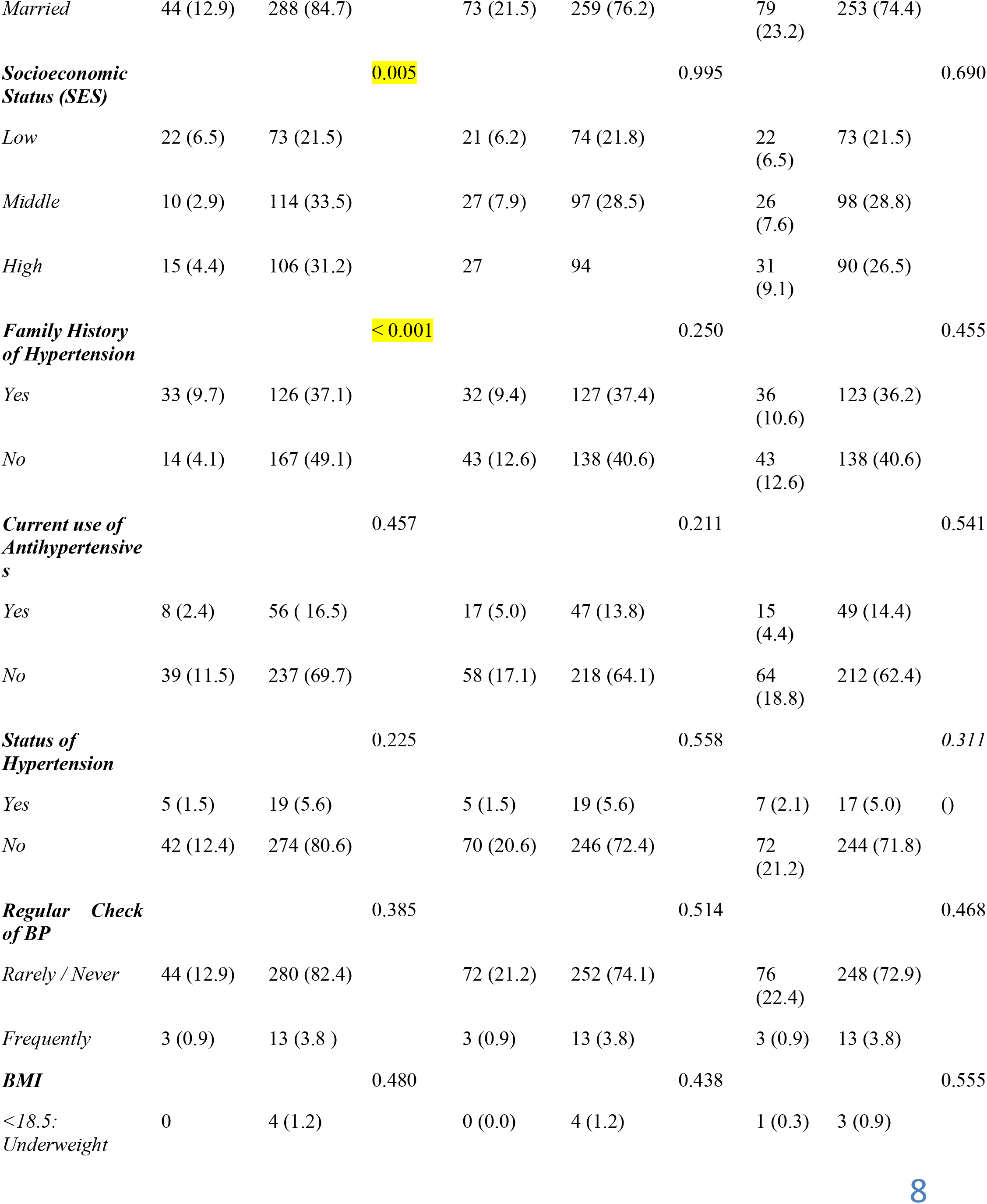
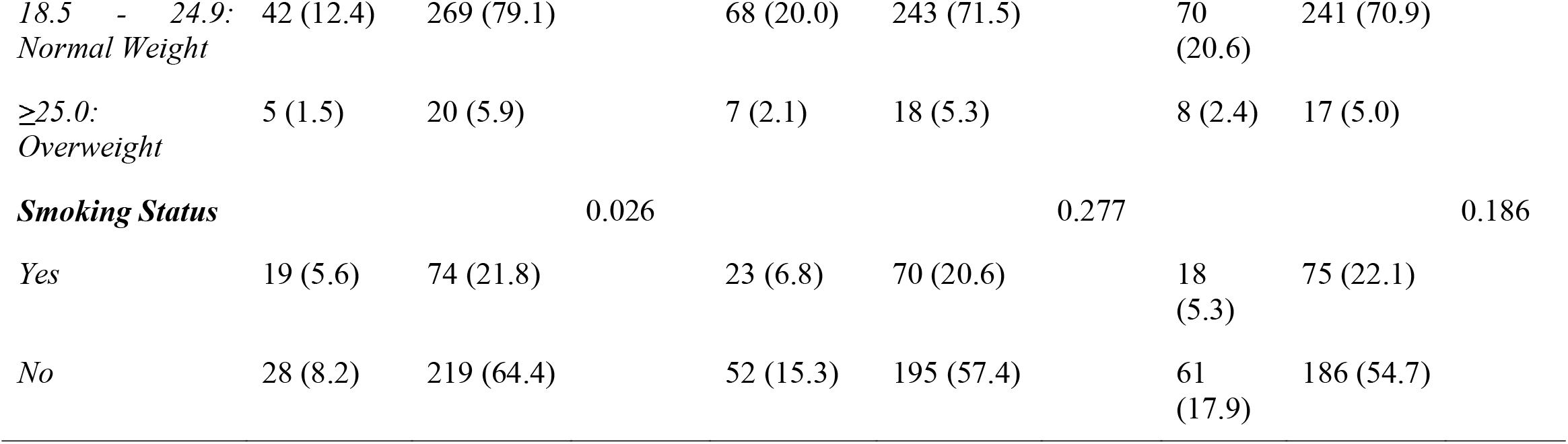
Chi-square test of socio-demographic factors associated with KAP.

Age was not significant in the difference of the KAP levels across brackets, p > 0.05. For instance, the age bracket 20–29 represented 3.8% for the correct knowledge and 31.8% incorrect knowledge. Other age brackets showed similar distributions; therefore, age was not a significant influential factor on KAP.

In the majority, knowledge was wrong and attitude negative, and practice poor among both males and females. There was no statistical difference by gender on these aspects, as p-value is greater than 0.05. Among males, correct knowledge was identified in 13.5% whereas that among females is only 0.3%.

Although individuals with higher education levels had relatively better KAP scores, the associations were not statistically significant (p > 0.05). Higher secondary education or above was associated with somewhat improved knowledge, 5.9% correct knowledge as compared to 0% of participants with primary education.

It means marital status does not have any significance on the level of KAP. Most of the married respondents had wrong knowledge 84.7%, negative attitude 76.2%, poor practice 74.4%.

Socioeconomic status was significantly associated with knowledge, p = 0.005, since higher SES was related to better knowledge scores. On the other hand, the same did not relate significantly to attitude, p = 0.995, and practices, p = 0.690.

Thus, there is a significant association between family history of hypertension and knowledge p < 0.001. Overall, subjects with a family medical history of hypertension had better knowledge scores 9.7% correct knowledge compared to those without a history 4.1%. However, family history did not influence either the attitude or practices significantly.

Other variables such as the use of anti-hypertensive medication at present, status of hypertension, frequency of blood pressure measurements, body mass index, and smoking status did not significantly affect the KAP scores. Smoking status marginally affected knowledge with a p-value of 0.026; however, it did not affect attitudes and practices. Non-smokers demonstrated a little better knowledge and attitude compared to smokers.

## Discussion

This has therefore brought to light significant deficits in KAP about hypertension that are necessary to bring into practice for better management of hypertension and health outcomes. This is in consistency with findings throughout studies in Bangladesh, Afghanistan, Ghana, and other LMICs where highly male-dominant, relatively well-educated samples showed poor mastery of basic concepts of hypertension. Furthermore, it was noted that only 13.8% of respondents had appropriate knowledge about the condition, especially with regards to necessary lifestyle behaviors such as weight control, salt reduction, and regular physical activity, long cited as paramount in the management and prevention of hypertension. The observed vast knowledge gap also implies that the formally educated population remains largely ignorant about the disease, either because of poor access to health information or due to lack of adequate dissemination through health education programs already in place. The findings point to the need for immediate tailor-made educational interventions, addressing these very knowledge gaps and providing information that is usable by the general population.

There was no difference in attitudes regarding management of hypertension. A whopping 77.9% were either unwilling or skeptical about preventive measures such as regular blood pressure monitoring, lifestyle modification, and drug compliance. This indifference to prevention aligns with the cultural health-seeking behavior that has been reported in different regions and is partly influenced by the lack of understanding of the chronic and usually asymptomatic nature of hypertension and its complications. A review of studies conducted in Iran and Ethiopia, among other LMICs, indicates that such preventive attitudes regarding the management of hypertension are influenced by cultural beliefs and norms, as well as a limited understanding of the grave health consequences of unmanaged hypertension, including stroke, kidney failure, and heart disease. Such negative attitudes will shift only with tailored public health campaigns that emphasize the long-term risks associated with hypertension and the benefits of proactive management. They should be culturally relevant and employ forms of communication that are local to them, including testimonials or other relatable illustrations to make the risks of hypertension relevant and more convincing to the audience.

Further still, the study reported on health practices, indicating that a meager 76.8% of respondents did not practice recommended health behaviors in the form of regular physical activity and dietary changes, and checking one’s blood pressure regularly. This low practice is in tandem with the trend from studies in countries such as Ghana and Ethiopia, where healthy practices were reported to be poorly adhered to due to a poor KAP score. These barriers may emanate from a variety of factors, including time constraints, lack of accessible resources, or inadequate health services that support or facilitate these practices. For example, busy work schedules or limited access to exercise facilities may impede regular physical activity, while financial constraints may affect one’s ability to afford healthier dietary choices. Such obstacles, of course, call for comprehensive interventions aimed at the socio-economic and logistic barriers to such health-enhancing activities. Such programs may include on-site wellness activities, community-based health promotion initiatives, and low-cost health care services-all tailored to give real-life support to fostering positive behavior.

This socio-demographic analysis of the study revealed that age, gender, and education level did not have much influence on the KAP score, indicating that knowledge and attitude gaps are fairly consistent across the demographic groupings. In the same period, some of the variables like higher socio-economic status and family history of hypertension were found to be positively associated with better knowledge scores. The finding agrees with studies from countries such as Egypt and Lebanon that stated that the socio-economic advantages along with personal exposure to hypertension increase the levels of awareness. Chances are that members of a high socio-economic class will have better access to health facilities, reading materials, and avenues to seek preventive care. Some of them may have family medical histories of hypertension, but they would have grown up hearing about how to manage the condition or observing what was happening around family members as a way to base their knowledge and practices on. These insights show that any future public health intervention needs to be equitable and reduce socio-economic disparities in health education and resource access. Moreover, awareness programs oriented toward the family and directed at households with a family history of hypertension could further solidify awareness and proactive management in high-risk groups.

This study has pointed out substantial gaps in KAP, which greatly hamper the proper management of hypertension among the respondents. Improvement in these gaps requires multiple strategies that can be directed toward enhancement in knowledge, fostering positive attitudes, and ensuring healthy practices across all socio-demographic spectrums. The programs of tailored health education, culturally sensitive campaigns for public health, and resource-supportive initiatives at workplaces and communities should collectively work toward raising a better-informed and more proactive population. These efforts have the potential to reduce overall hypertension burden and risks, resulting in improved health outcomes that might also lighten the future burden on healthcare systems.

## Conclusion

The present study has underlined the key knowledge gaps, attitudes, and practices among participants regarding hypertension management and pointed out serious barriers as far as prevention and control of the disease are concerned. The study further shows low awareness of basic concepts with regard to hypertension and lifestyle changes, along with negative attitude toward prevention targeted by educational interventions that include identified specific knowledge deficits and cultural attitudes. Generally poor adherence to recommended health practices is also indicative of socio-economic and logistical barriers in the light of adequate hypertension self-management.

This means interventions from this research should be multi-layered in creating health literacy across diverse demographic groups. Socio-economic realities and cultural contexts would also have to be tailor-made. It would be possible through equal opportunities in accessing health resources, community-based health programs, and workplace wellness initiatives for disseminating information and providing the necessary support for positive behavioral change. Such awareness campaigns involving the family may focus more on households in which cases of hypertension have been registered earlier, thereby ensuring better awareness and proactive management among high-risk persons. Each of these factors, if addressed, will further help decrease the prevalence of hypertension through public health strategy and reduce its chronic health consequences, hence improving health outcomes for the population and easing burdens on healthcare systems.

## Data Availability

Data Availability Statement:
All data generated or analyzed as part of this study have been comprehensively included in this published article. Any supplementary information files associated with this publication also contain relevant data, ensuring full transparency and accessibility for further analysis or reference.

https://drive.google.com/file/d/1gYNt1xyovE1ChkpZPHpk3JQf1sXuOcog/view?usp=drive_link

## Acknowledgment

*In this regard, we would like to extend our appreciation to all the employees of the beverage company who participated in this research*.

## Scientific Responsibility Statement

*The authors also affirmed that they bear full responsibility for the scientific contents of the article starting from the study design, the collection of data, the analysis, and interpretation of data, writing the article, some of the main ideas, or part of preparation and scientific peer review of the contents and the final approval of the article*.

## Animals and human rights declaration

*In this study, all procedures are conducted to determine the confidentiality or identify the patients according to the ethical standards of the institutional and/or national research committee, the declaration of Helsinki of 1964, and its amendments or similar guidelines. The authors of this article did not conduct any animal or human studies for this article*.

## Data Availability Statement

All data generated or analyzed as part of this study have been comprehensively included in this published article. Any supplementary information files associated with this publication also contain relevant data, ensuring full transparency and accessibility for further analysis or reference.

